## Supplementary file questionnaire contents for "UK osteopathic practice in 2019: a retrospective analysis of practice data"

**Questionnaire survey content**

Part A provided full information about the survey and obtained consent.

Part B included questions about:

- Osteopath's gender, age, and nationality
- Number of years of experience as an osteopath after initial graduation
- Practicing status
- Academic qualifications
- Working status as an osteopath (i.e. independent, employee)
- Location of practice (urban or rural)
- Practice environment (i.e. private practice, group practice with other osteopaths, interdisciplinary practice, clinic, hospital)

Part C included questions about:

- Description of patient and underlying condition:
- Month of encounter for selected patient
- Patient's age
- Patient's gender
- Primary reason for consultation
- Co-existing known conditions
- Duration of actual episode
- Type of onset of symptoms
- Localisation of symptoms
- Impact of symptoms on daily life
- Prior care for actual episode
- Any health professional who referred the patient

▪ Description of first encounter:

- Informed consent for examination and treatment
- Treatment techniques employed during first encounter
- Use of adjunct therapies
- Recommendation of self-management strategies
- Duration of first encounter

▪ Subsequent encounters:

- Number of encounters for primary reason for consultation
- Number of subsequent treatments (including adjunct therapies and self-management strategies)
- If applicable, description of referral to other health professional
